# A Risk Score to Predict Admission to Intensive Care Unit in Patients With COVID-19: The ABC-GOALS Score

**DOI:** 10.1101/2020.05.12.20099416

**Authors:** Juan M. Mejía-Vilet, Bertha M. Córdova-Sánchez, Dheni A. Fernández-Camargo, R. Angélica Méndez-Pérez, Luis E. Morales Buenrostro, Thierry Hernández-Gilsoul

**Author notes:** **Correspondence:** Thierry Hernández-Gilsoul, Head of the Emergency Department, Instituto Nacional de Ciencias Médicas y Nutrición Salvador Zubirán, Mexico City, Mexico, Vasco de Quiroga 15, Belisario Domínguez Sección XVI, Tlalpan, Mexico City, Mexico, 14080, ABC-GOALS score to predict ICU admission.

## Abstract

**Objectives:** COVID-19 pandemic poses a burden on hospital resources and intensive care unit (ICU) occupation. This study aimed to provide a scoring system that, assessed upon first-contact evaluation at the emergency department, predicts the need for ICU admission.

**Methods:** We prospectively assessed patients admitted to a COVID-19 reference center in Mexico City between March 16^th^ and May 21^st^, and split them into development and validation cohorts. Patients were segregated into a group that required admission to ICU, and a group that never required ICU admission and was discharged from hospitalization. By logistic regression, we constructed predictive models for ICU admission, including clinical, laboratory, and imaging findings from the emergency department evaluation. The ABC-GOALS score was created by assigning values to the weighted odd ratios. The score was compared to other COVID-19 and pneumonia scores through the area under the curve (AUC).

**Results:** We included 569 patients divided into development (n=329) and validation (n=240) cohorts. One-hundred-fifteen patients from each cohort required admission to ICU. The clinical model (ABC-GOALS_c_) included sex, obesity, the Charlson comorbidity index, dyspnea, arterial pressure, and respiratory rate at triage evaluation. The clinical plus laboratory model (ABC-GOALS_cl_) added serum albumin, glucose, lactate dehydrogenase, and S/F ratio to the clinical model. The model that included imaging (ABC-GOALS_clx_) added the CT scan finding of >50% lung involvement. The model AUC were 0.79 (95%CI 0.74-0.83) and 0.77 (95%CI 0.71-0.83), 0.86 (95%CI 0.82-0.90) and 0.87 (95%CI 0.83-0.92), 0.88 (95%CI 0.84-0.92) and 0.86 (95%CI 0.81-0.90) for the clinical, laboratory and imaging models in the development and validation cohorts, respectively. The ABC-GOALS_cl_ and ABC-GOALS_clx_ scores outperformed other COVID-19 and pneumonia-specific scores.

**Conclusion:** The ABC-GOALS score is a tool to evaluate patients with COVID-19 at admission to the emergency department, which allows to timely predict their risk of admission to an ICU.

## INTRODUCTION

The presence of a new severe acute respiratory syndrome coronavirus 2 (SARS-CoV-2) was first reported in China and has subsequently spread to all regions of the world, straining the health systems of many countries^1^. Viral pneumonia associated with SARS-CoV-2 has been officially denominated as coronavirus disease 2019 (COVID-19)^2^.

Approximately 5 to 33% of patients with COVID-19 pneumonia will be admitted to an intensive care unit (ICU)^3–5^. Previous studies have identified several risk factors associated with a severe course of COVID-19 pneumonia and its progression to acute respiratory distress syndrome ^6,7^. These risk factors may be categorized into patient characteristics obtained through the medical interview (e.g. age, comorbidities, symptoms)^6–11^; vital signs obtained from triage evaluation (e.g. respiratory rate, arterial pressure)^12,13^; laboratory abnormalities including inflammatory, coagulation and organ-specific studies (e.g. lactate dehydrogenase, D-dimer, fibrinogen, cardiac troponins, liver function tests, among others) ^6,7,9–11,14^; and lung imaging findings (e.g. number of affected lobes, estimated pneumonia extension) ^15,16^.

In several countries, including Mexico, hospital reconversion and temporary care centers have been implemented to cope with a large number of COVID-19 patients ^17^ However, many of these centers are prepared to care for patients with supplemental oxygen requirements in general wards, but just a few patients who require admission to an intensive care unit (ICU) and mechanical ventilation.

The study aimed to derive and validate simple risk prediction models to anticipate the need for admission to an ICU. These models include clinical, laboratory, and image findings obtained at the first-contact evaluation to aid triage, decision-making, and timely referral to maintain healthcare system capacity.

## METHODS

This is a prospective observational cohort study. All consecutive adult (>18 years) patients hospitalized at Instituto Nacional de Ciencias Médicas y Nutrición Salvador Zubirán, a referral center for COVID-19 patients at Mexico City, were evaluated for this analysis. The study was approved by the local Research and Ethics Board *(CAI-3368-20-20-1)* that waived the use of a written informed consent form due to the study’s nature.

All patients hospitalized between March 16^th^ and April 30^th^ were assigned to the development cohort, and patients hospitalized between May 1^st^ and May 21^th^ were assigned to the validation cohort. The study population was segregated into two groups: 1) patients who required admission to an ICU at any time during their hospitalization, and 2) patients admitted to hospital general wards who were discharged from hospitalization without ever been considered for admission to an ICU. Patients were censored for each group once they were admitted to the ICU or by the date of discharge from hospitalization to home, respectively. All patients who remained hospitalized in general wards by the end of the study were not included in this analysis as it was considered that their risk for ICU admission was still active.

All patients had COVID-19 pneumonia diagnosed by chest computed tomography (CT), and SARS-CoV-2 infection confirmed in respiratory specimens using real-time reverse-transcriptase polymerase chain reaction (RT-PCR) assay by local testing and confirmed at a central laboratory. Patients were considered positive if the initial test results were positive, or if it was negative but repeat testing was positive.

The following variables were obtained by the time of the triage and emergency department (ED) evaluation: demographic variables, previous medical and medication history including smoking; symptoms, physical examination including weight, height, and vital signs; laboratory evaluation including arterial blood gas analysis, inflammatory biomarkers, troponin-I levels, complete blood count and blood chemistries; and chest CT-scan findings. All patients underwent chest CT-scans that were evaluated by experienced specialists; lung involvement was semi-quantitatively classified as mild (<20%), moderate (20-50%), or severe (>50%).

For each patient, we calculated the Charlson comorbidity index (an index that predicts 10-year survival in patients with multiple comorbidities) ^18^, National Early Warning Score (NEWS) 2 ^19^, Sequential Organ Failure Assessment (SOFA) score ^20^, CURB-65 score for pneumonia severity ^21^, the MuLBSTA score for viral pneumonia mortality ^22^, the ROX index to predict the risk of intubation ^23^, the CALL score model for prediction of progression risk in COVID-19 ^24^ and the COVID-GRAM score for the risk of critical illness in COVID-19^25^. All scores were calculated at admission to the ED. Although some of these scores were not derived to evaluate the risk of admission to an ICU, we evaluated their value if repurposed for this end.

The need for admission to critical care was determined by the medical team in charge of the patient and included the need for mechanical ventilation or high-dose vasopressors. Patients hospitalized in medical wards were treated with supplementary oxygen (nasal cannula or non-rebreathing oxygen mask) but were never considered for intubation and ICU admission. All patients in the latter group were censored once discharged from hospitalization to home after improvement. Clinical outcomes for patients admitted to ICU were monitored up to May 25^th’^ 2020.

### Statistical analysis

The distribution of continuous variables was evaluated by the Kolmogorov-Smirnov test. Variables are described as number (relative frequency) or median (interquartile range [IQR]) as appropriate. Characteristics at admission between study groups were compared by the Mann-Whitney U test. There were less than 2% missing values for all variables. In the case of missing data, variables were imputed by using multiple imputations ^26^

For score derivation, all variables were evaluated by bivariate logistic regression analysis. All variables with p-values <0.05 were considered for the multivariate logistic regression analysis. The best logistic regression analysis model was constructed by the forward stepwise selection method using maximum likelihood estimation and r-square values. Three models were constructed: a first model including only clinical variables (ABC-GOALS_c_), a second model that included clinical and laboratory variables (ABC-GOALS_cl_), and a third model that included clinical, laboratory, and x-ray findings (ABC-GOALS_clx_). The predictive performance of each model was measured by the concordance index (C-index) and internal calibration was evaluated by 1000 bootstrap samples. The goodness of fit was evaluated by the Hosmer-Lemeshow test. To create the final scores, points were assigned by the weighted odd-ratios and approached to the closest integer for each model.

We prospectively validated the ABC-GOALS scores in an independent validation cohort. The performance of the derived scores as well as for all other scores determined at admission to predict the risk of hospitalization into an ICU were assessed by receiver operating characteristic (ROC) curves and their 95% confidence intervals.

All analyses were performed with SPSS version 24.0 (IBM, Armonk, NY, USA) and GraphPad Prism 6.0 (GraphPad Software, San Diego, CA, USA).

## RESULTS

### Patient characteristics at admission

A total of 705 patients with COVID-19 pneumonia and SARS-CoV-2 positive test were hospitalized during the study period. Eighty-three patients who remained hospitalized in general wards, 28 patients referred to a convalescence center, 22 early referred to other institutions, and three patients who were discharged by discharge against medical advice were excluded from the analysis. The development cohort included 329 patients and the validation cohort 240 patients (Figure 1).

**Figure 1.**
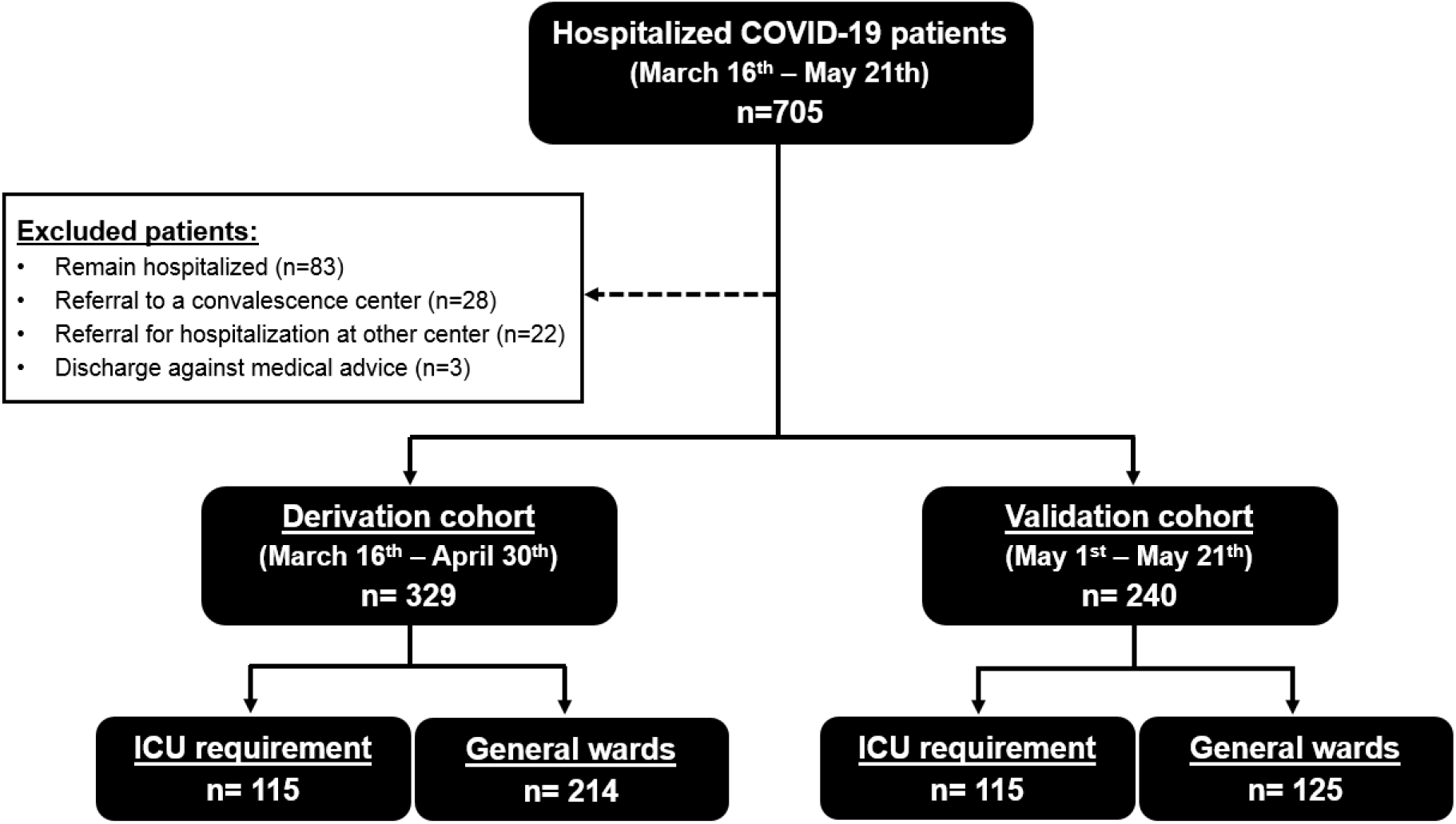
Flow diagram showing all subjects included in the study.

### Characteristics of the development cohort

The development cohort comprised 115 patients that required ICU admission and 214 patients that had been hospitalized and discharged from general wards. The median age of the derivation cohort was 49 years (IQR 41-60), 211 (64%) subjects were male, with a median Charlson comorbidity index of 1 point (IQR 0-2). There were 100 (30%) subjects with no previous comorbidity. The median days from the start of symptoms to the evaluation at the ER were 7 days (IQR 5-10). All patients received supplementary oxygen. For the group admitted to ICU, the median time from ER admission to ICU admission was 2 days (IQR 0-3 days, range 0 to 12). The median length of hospitalization for the group hospitalized in general wards was 6 days (IQR 4-9 days, range 1 to 20). Several differences in the admission variables were observed between patients who required critical care and those hospitalized in general wards and are described in Table 1 and Supplementary Table 1.

**Table 1.**
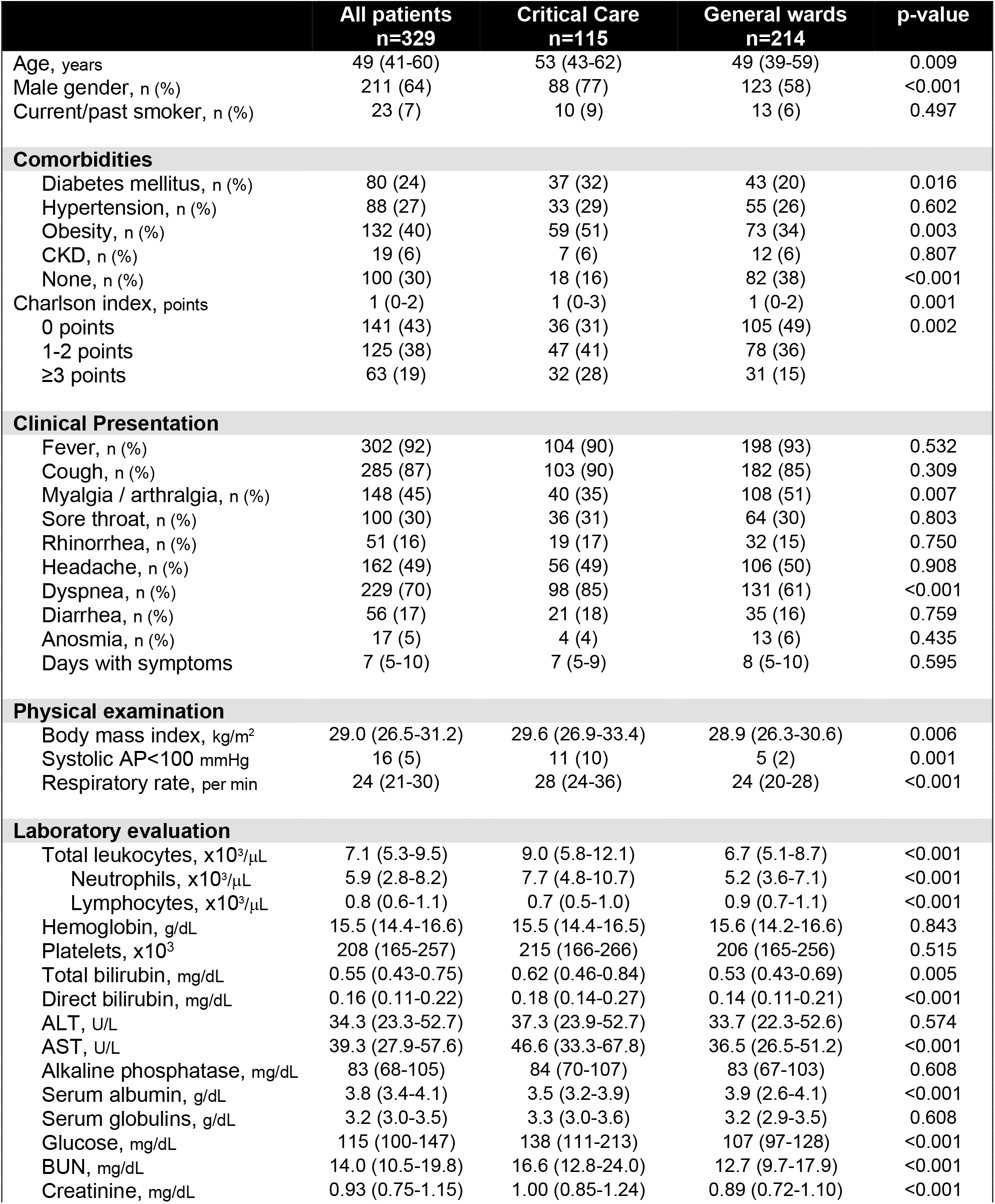
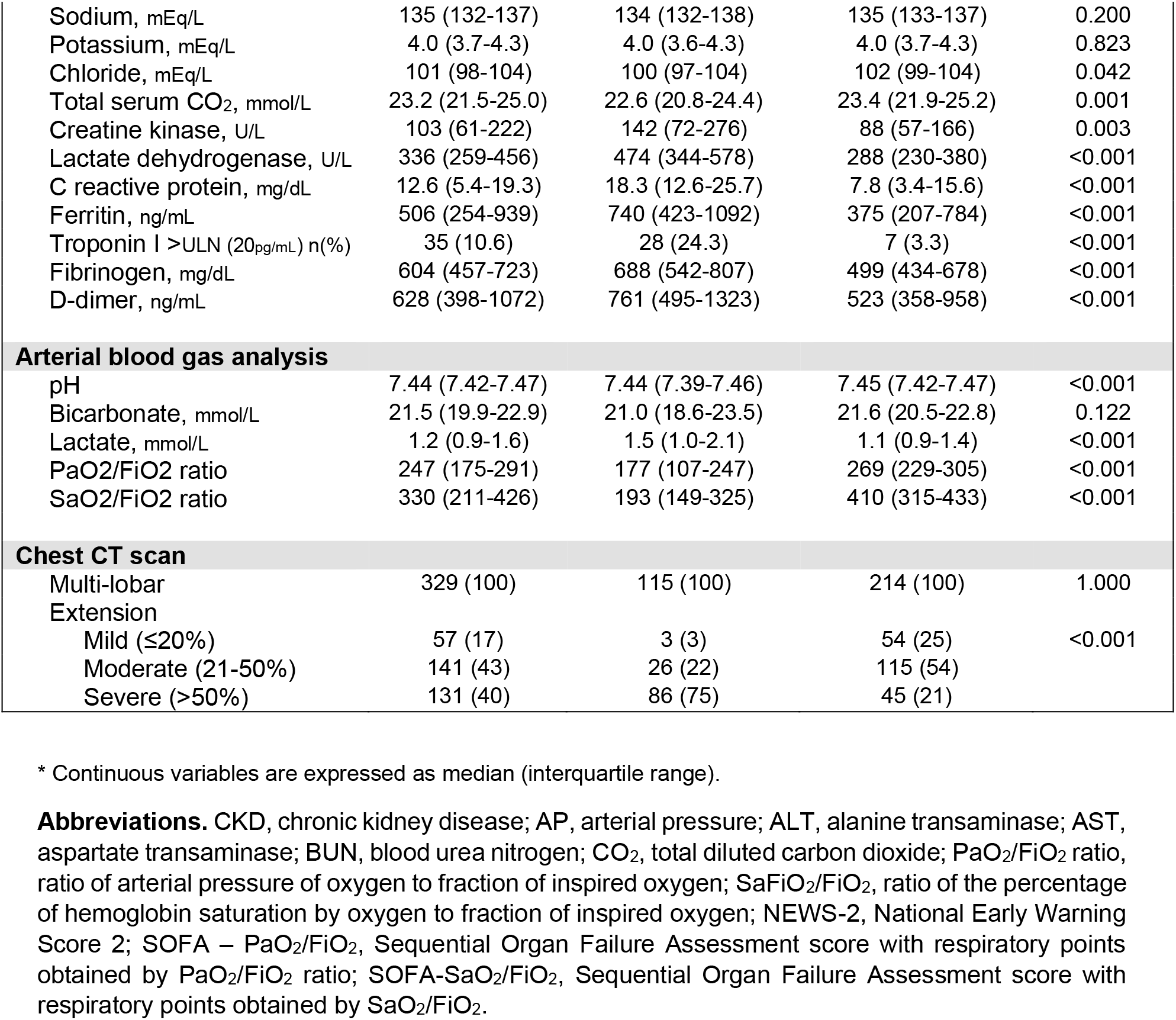
Characteristics of patients in the development cohort at first evaluation in the emergency department.

### Factors associated with the need for admission to a critical care unit

An extended table with all the results obtained from the bivariate logistic regression analysis is shown in Supplementary Table 2. The best predictive models derived from multivariable logistic regression analysis are shown in Table 2. The clinical model included: male sex, the Charlson comorbidity index, obesity (BMI≥30kg/m^2^, not included in the Charlson index), referred dyspnea, respiratory rate, and systolic arterial pressure at the triage or ER evaluation. The clinical plus laboratory model included the same clinical variables plus serum albumin <3.5g/dL, lactate dehydrogenase above the upper limit of normal, and the hemoglobin oxygen saturation to the fraction of inspired oxygen ratio <300 (S/F ratio). The clinical plus laboratory model included all previous variables except for referred dyspnea and serum albumin and added reported lung involvement >50% in the lung CT scan. The c-statistics for clinical, clinical plus laboratory and clinical plus laboratory plus x-ray models were 0.79, 0.87, and 0.88, respectively.

**Table 2.** Models to predict the risk of admission to an intensive care unit

|  | OR | Bivariate<br>95%CI | p-value | OR | Multivariate<br>95%CI | p-value |
| --- | --- | --- | --- | --- | --- | --- |
| **Model 1: clinical** |  |  |  |  |  |  |
| Male gender | 2.41 | 1.45-4.01 | 0.001 | 1.82 | 1.01-3.30 | 0.049 |
| Obesity, BMI≥30kg/m<sup>2</sup> | 2.04 | 1.28-3.23 | 0.003 | 2.56 | 1.56-4.55 | 0.001 |
| Dyspnea | 3.65 | 2.04-6.55 | <0.001 | 2.39 | 1.22-4.67 | 0.022 |
| SBP<100mmHg | 4.42 | 1.50-13.06 | 0.007 | 5.10 | 1.37-18.9 | 0.015 |
| Respiratory rate |  |  |  |  |  |  |
| <24 per min | 1.00 | reference | reference | 1.00 | reference | reference |
| 24-28 per min | 2.253 | 1.223-4.150 | 0.009 | 2.12 | 1.08-4.17 | 0.029 |
| >28 per min | 8.063 | 4.279-15.19 | <0.001 | 5.14 | 2.50-10.7 | <0.001 |
| Charlson index |  |  |  |  |  |  |
| 0 points | 1.00 | reference | reference | 1.00 | reference | reference |
| 1-2 points | 1.76 | 1.04-2.97 | 0.035 | 2.33 | 1.28-4.27 | 0.006 |
| ≥3 points | 3.01 | 1.62-5.61 | 0.001 | 4.96 | 2.32-10.6 | <0.001 |
| **Model 2: clinical + laboratory** |  |  |  |  |  |  |
| Male gender | 2.41 | 1.45-4.01 | 0.001 | 2.48 | 1.28-4.81 | 0.007 |
| Obesity, BMI≥30kg/m<sup>2</sup> | 2.04 | 1.28-3.23 | 0.003 | 2.97 | 1.59-5.55 | 0.001 |
| Dyspnea | 3.65 | 2.04-6.55 | <0.001 | 2.24 | 1.08-4.61 | 0.029 |
| Charlson index ≥3 | 2.28 | 1.30-3.98 | 0.004 | 2.84 | 1.31-6.15 | 0.008 |
| SBP<100mmHg | 4.42 | 1.50-13.06 | 0.007 | 5.71 | 1.31-24.9 | 0.020 |
| Glucose >200g/dL | 3.10 | 1.67-5.74 | <0.001 | 3.00 | 1.35-6.67 | 0.007 |
| Albumin <3.5g/dL | 4.15 | 2.52-6.83 | <0.001 | 2.13 | 1.14-3.99 | 0.018 |
| LDH>ULN | 8.55 | 4.11-17.79 | <0.001 | 3.39 | 1.47-7.79 | 0.004 |
| S/F ratio<300 | 8.48 | 5.07-14.19 | <0.001 | 5.41 | 2.93-9.97 | <0.001 |
| **Model 3: clinical + laboratory + image** |  |  |  |  |  |  |
| Male gender | 2.41 | 1.45-4.01 | 0.001 | 2.14 | 1.09-4.18 | 0.026 |
| Obesity, BMI≥30kg/m<sup>2</sup> | 2.04 | 1.28-3.23 | 0.003 | 2.58 | 1.37-4.85 | 0.003 |
| Charlson index ≥3 | 2.28 | 1.30-3.98 | 0.004 | 2.90 | 1.32-6.37 | 0.008 |
| SBP<100mmHg | 4.42 | 1.50-13.06 | 0.007 | 5.15 | 1.12-23.6 | 0.035 |
| Glucose >200mg/dL | 3.10 | 1.67-5.74 | <0.001 | 2.64 | 1.14-6.12 | 0.023 |
| LDH>ULN | 8.55 | 4.11-17.79 | <0.001 | 2.83 | 1.20-6.66 | 0.017 |
| S/F ratio<300 | 8.48 | 5.07-14.19 | <0.001 | 4.43 | 2.35-8.35 | <0.001 |
| CT lung involvement >50% | 11.1 | 6.53-18.99 | <0.001 | 4.62 | 2.47-8.64 | <0.001 |

**Abbreviations.** OR, odds ratio; 95%CI, 95% confidence intervals; SBP, systolic blood pressure; LDH, lactate dehydrogenase serum levels; S/F ratio, ratio of percentage of hemoglobin saturation to fraction of inspired oxygen; CT, computed tomography.

### Derivation of the ABC-GOALS scores

A predictive point score was constructed based on the weighted OR’s from each logistic regression model. The derived scores were defined as the ABC-GOALS (Arterial pressure, Breathlessness, Charlson, Glucose, Obesity, Albumin, LDH and S/F ratio) and were labeled as clinical (ABC-GOALS_c_), clinical plus laboratory (ABC-GOALS_cl_) and clinical plus laboratory plus x-ray (ABC-GOALS_clx_) scores (Figure 2).

**Figure 2.**
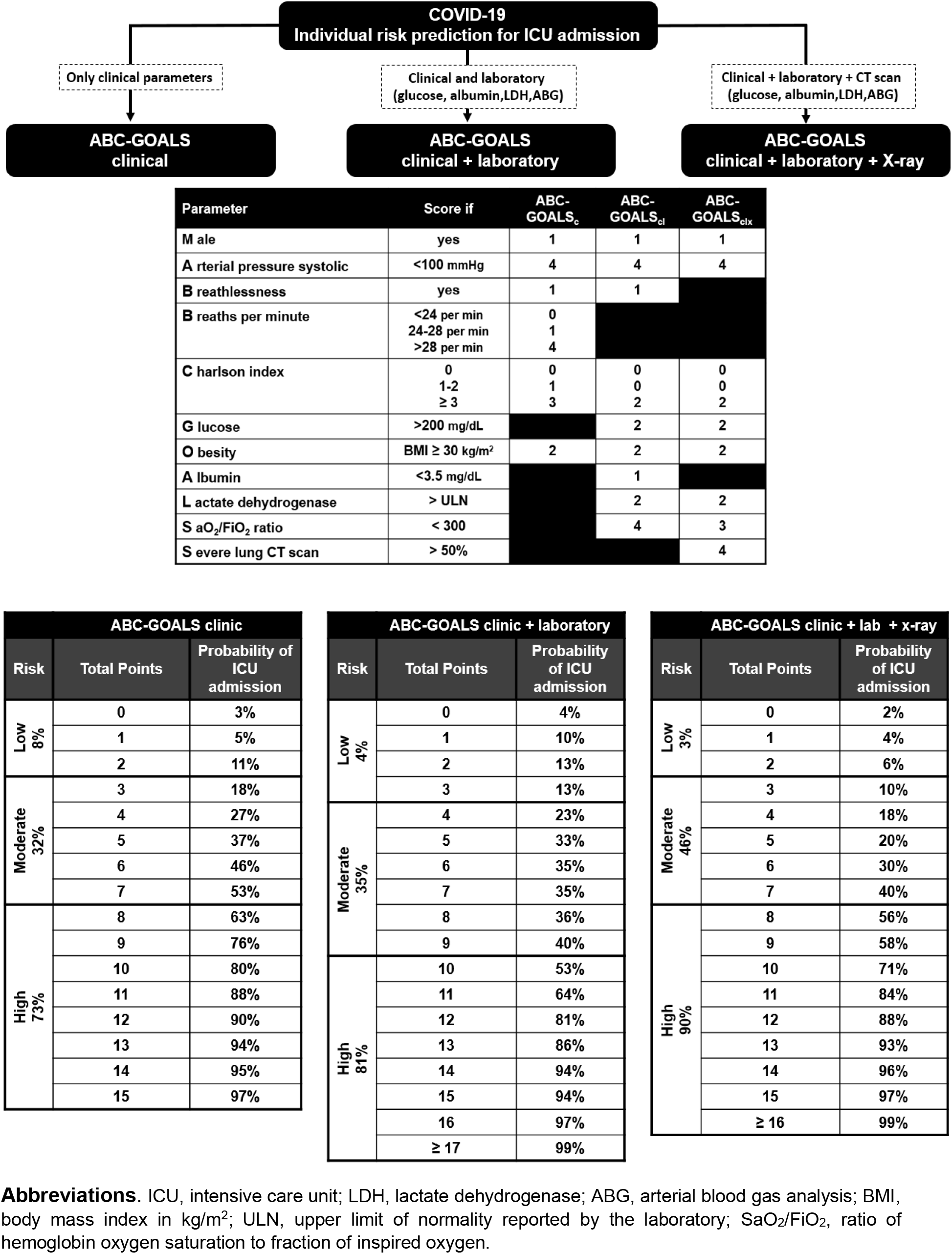
Guide to ABC-GOALS score calculation.

All three ABC-GOALS score variations demonstrated good accuracy in estimating the risk of admission to an ICU, with an area under the curve of 0.79 (95%CI 0.74-0.83), 0.86 (95%CI 0.82-0.90) and 0.88 (95%CI 0.84-0.92) for the ABC-GOALS_c_, ABC-GOALS_cl_ and ABC-GOALS_clx_ scores, respectively (Figure 3). Calibration plots showed good agreement between the estimated and observed scores (Supplementary Figure 1).

**Figure 3.**
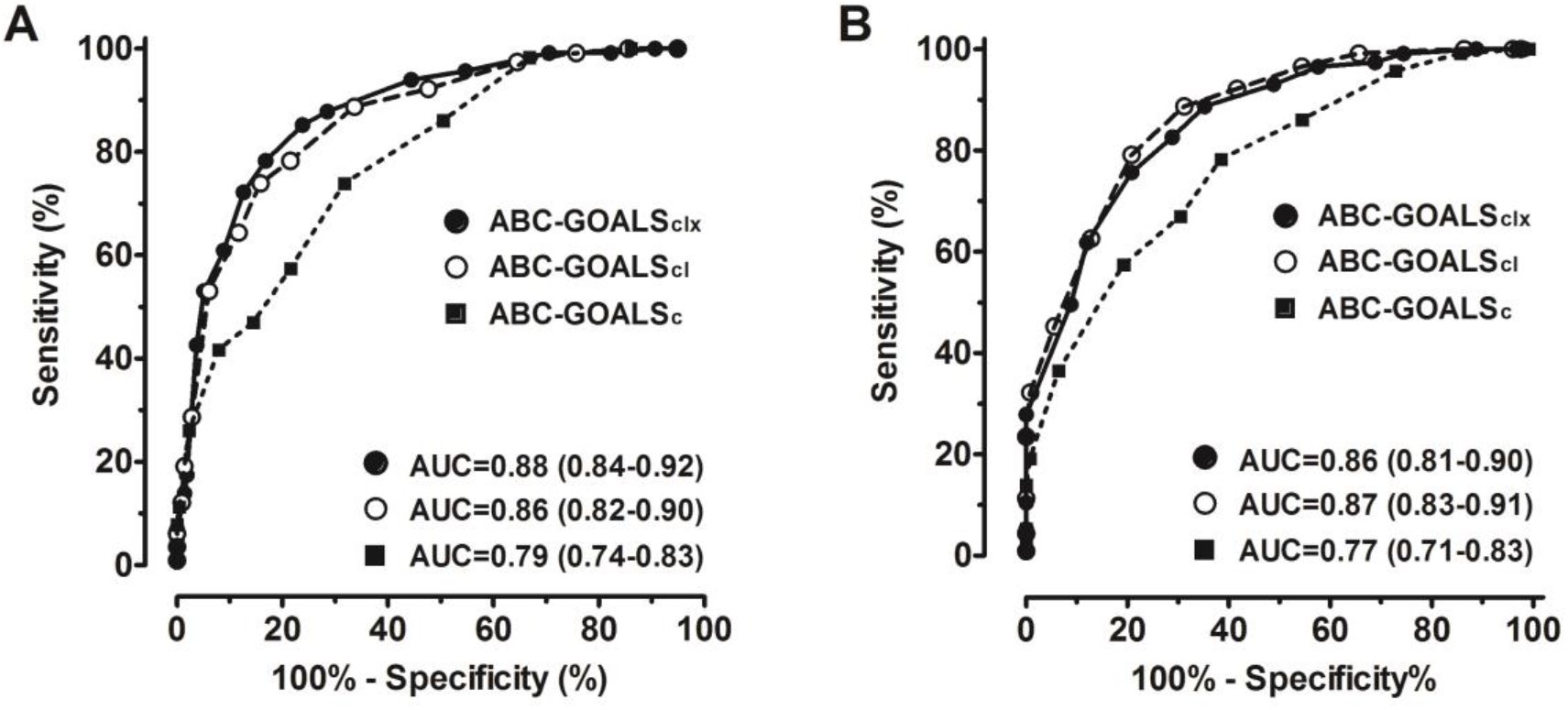
Receiver operating curves for the ABC-GOALS scores in the development (A) and validation (B) cohorts.

By segregating the scores into three levels of risk of admission to ICU, the ABC-GOALS_c_ classified patients into low- (0-2 points, mean risk 8%, 95%CI 7-9%), moderate- (3-7 points, mean risk 32%, 95%CI 30-34%) and high-risk (≥8 points, mean risk 73%, 95%CI 70-75%) of admission ICU. The respective groups for the ABC-GOALS_cl_ were low- (0-3 points, mean risk 4%, 95%CI 3-4%), moderate- (4-9 points, mean risk 30%, 95%CI 27-32%) and high-risk (≥10 points, mean risk 80%, 95%CI 78-82%) for ICU admission. Finally, the groups for the ABC-GOALS_clx_ were low- (0-4 points, mean risk 6%, 95%CI 5-6%), moderate- (5-9 points, mean 32%, 95%CI 29-34%) and high-risk (≥10 points, mean 80%, 95%CI 78-82%) for ICU admission (Figure 2). The sensitivity and specificity for selected cutoffs of these scores are shown in Table 3.

**Table 3.** Predictive performance for admission to ICU of all the evaluated scores.

| Score | Area under the curve |  |
| --- | --- | --- |
|  | Development cohort | Validation cohort |
| **ABC-GOALS<sub>c</sub>** | 0.79<br>(0.74-0.83) | 0.77<br>(0.71-0.83) |
| **ABC-GOALS<sub>cl</sub>** | 0.86<br>(0.82-0.90) | 0.87<br>(0.83-0.92) |
| **ABC-GOALS<sub>clx</sub>** | 0.88<br>(0.84-0.92) | 0.86<br>(0.81-0.90) |
| **COVID-GRAM** | 0.78<br>(0.73-0.83) | 0.77<br>(0.72-0.83) |
| **CALL score** | 0.72<br>(0.67-0.78) | 0.69<br>(0.63-0.76) |
| **SOFA** | 0.77<br>(0.71-0.82) | 0.73<br>(0.66-0.79) |
| **NEWS-2** | 0.76<br>(0.70-0.81) | 0.64<br>(0.57-0.71) |
| **CURB-65** | 0.71<br>(0.65-0.77) | 0.74<br>(0.67-0.80) |
| **MuLBSTA** | 0.63<br>(0.57-0.70) | 0.58<br>(0.50-0.65) |
| **ROX index** | 0.81<br>(0.76-0.86) | 0.84<br>(0.80-0.90) |

\* All scores were obtained at admission to the emergency room. SOFA respiratory points were assigned based on the S/F ratio.
**Abbreviations.** ABC-GOALS<sub>c</sub>, score including only clinical variables; ABC-GOALS<sub>cl</sub>, score including both clinical and laboratory variables; ABC-GOALS<sub>clx</sub>, score including clinical, laboratory and x-ray findings; NEWS-2, National Early Warning Score version 2; CALL, comorbidity, age, lymphocyte, LDH score for predicting progression of COVID-19; SOFA, Sequential Organ Failure Assessment score; CURB-65, confusion, urea, respiratory rate, blood pressure, age>65 years score for risk of mortality from pneumonia; MuLBSTA, Multi-lobar infiltrate, Lymphocyte count, Bacterial co-infection, Smoking history, hypertension, Age>65 years score for mortality from viral pneumonia; ROX index, S/F ratio divided by the respiratory rate to predict the risk of intubation in hypoxemic respiratory failure.

The ABC-GOALS variants outperformed other scores that were repurposed for the prediction of admission to ICU, based on the area under the curve values (Table 3). As previously stated, most of these scores were not created to predict this outcome. For practicality, the three ABC-GOALS scores were implemented into an application for mobile devices.

### Validation of the ABC-GOALS scores

The ABC-GOALS scores were prospectively validated in consecutive patients included into the validation cohort. The characteristics of the patients included in this cohort are shown in Supplementary Table 3. The area under the curve for each score was 0.77 (95%CI 0.71-0.83), 0.87 (95%CI 0.83-0.91), and 0.86 (95%CI 0.81-0.90) for the ABC-GOALS_c_, ABC-GOALS_cl_, and ABC-GOALS_clx_, respectively (Figure 3). The ABC-GOALS scores’ predictive performance was also superior to other scores in the validation cohort (Table 3).

### Outcomes

By the end of this study (May 24^th^), all patients hospitalized in general wards in the development and validation cohorts were discharged to home after improvement. Three patients (0.8%) were readmitted and discharged again without admission to ICU during the study period. From all patients admitted to the ICU in the development cohort (n=115), 63 (55%) have died, 37 (32%) have been discharged to their homes after improvement, and 15 (13%) remain hospitalized.

## DISCUSSION

In this study, we derived and validated a prognostic score that, evaluated upon patient admission to the ED, helps predict the probability of admission to ICU during patient hospitalization. This tool may prove useful for patient assessment at first-contact evaluation and to timely refer patients to other units in case of a lack of or overcrowding of local ICUs. The ABC-GOALS scores predicted the need for admission to an ICU with greater accuracy than other reported COVID-19 prognostic scores, as well as other scores that were created for the prediction of other outcomes in sepsis or pneumonia, such as mortality.

To date, more than 4 million SARS-CoV-2 infections have been reported worldwide with more than 300,000 total deaths^27^. The fast spread of the disease ^28^ and the large number of patients admitted to hospitals has strained and overwhelmed local health systems. Among hospitalized patients, 5 to 33% will require admission to an ICU and 75% to 100% of them will require support with mechanical ventilation ^3–5,8^. An effective evaluation of patients with severe disease may be critical to maintain the healthcare system functional for as long as possible.

The ABC-GOALS scores summarize many of the previously reported factors that have been shown to independently associate with severe COVID-19 disease. We found that age and comorbidities predictive performance is enhanced when integrated into the Charlson comorbidity index. It has been reported in other studies that there may be interactions between age and some comorbidities such as diabetes^29^. Also, the Charlson comorbidity index offers the advantage of integrating several comorbidities into one score that has been previously associated with survival ^18^ Among clinical symptoms, dyspnea or shortness of breath is the most consistently reported symptom associated with COVID-19 severity and mortality ^6,9,12,30^ and was independently associated with ICU admission in this study. Obesity is a long-recognized factor for severe pulmonary infections ^31^ and has been consistently associated with adverse outcomes in COVID-19 ^32^ Low systolic arterial pressure and respiratory rate at triage evaluation complete the clinical ABC-GOALS.

Among the three derived scores in this study, the clinical ABC-GOALS has an inferior performance and probably equivalent to other early warning scores. It was designed to provide a tool for areas without timely access to laboratory results. However, the clinical score remains to be externally validated, as some reported models based exclusively on demographic (but not physical examination) information have been shown to decrease their performance after external validation^12^.

The scores integrating clinical with laboratory (ABC-GOALS_cl_) and imaging (ABC-GOALS_clx_) findings seem to perform better for prediction of admission to an ICU. Both models include values of glucose, lactate dehydrogenase, and oxygenation at admission that have been previously reported to be associated with prognosis in COVID-19^9,11,33,34^. Lactate dehydrogenase has been previously integrated into a predictive model (*see below*). In contrast to other laboratory tests that may increase during the progression of the disease, such as D-dimer and cardiac troponin levels ^35,36^, LDH has proved to be the most robust laboratory predictor for admission to ICU when evaluated at first-contact. Glucose and oxygenation were also included in these models. Oxygenation, reflected by the S/F ratio, probably reflects the disease extension along with the imaging findings. It is worth noting that the model that included imaging findings (ABC-GOALS_clx_) had an equivalent performance to the clinical and laboratory model, therefore, it may not be necessary to perform imaging in a patient who is planned to be referred based on the clinical plus laboratory model (ABC-GOALS_cl_).

We evaluated two scoring models that have been derived to predict the risk of COVID-19 progression: the CALL score^24^ and the COVID-GRAM score^25^. Both were derived from Chinese populations. As shown, both scores had lower performance than the ABC-GOALS scores in our population. The CALL score^24^ includes the presence of any comorbidity, age, lymphocyte count, and LDH levels. Most patients in our study presented with lymphocyte count below the cut-off set in the CALL score, therefore reducing the predictive value of the score and of this specific parameter. For the COVID-GRAM score^25^, it includes X-ray abnormalities, which were present in all of our patients by lung CT-scan. It also includes rare manifestations of COVID-19 that were barely documented in our population, such as hemoptysis and unconsciousness. Both, the CALL and COVID-GRAM scores, were derived from populations with a lower risk of critical care admission than our population, as 19.2% and 8.2% of the patients developed critical illness, respectively, compared to 34.9% and 47.9% in our development and validation cohorts, respectively. Thus, specific items included in the scores plus the derivation from a population with a lower risk for ICU admission may explain the lower performance of these scores in our population.

We showed that repurposing of other scores employed in pneumonia may not perform adequately to predict the need for admission to ICU in COVID-19 pneumonia. For example, the MuLBSTA score ^22^ has been validated for mortality prediction in viral pneumonia but may have a lower performance in COVID-19 pneumonia. This score includes the number of affected lobes by viral pneumonia, however, all patients in our study showed multi-lobar lung infiltrates reducing the performance of this parameter. Other variables included in this score, such as bacterial coinfection, may not perform well when evaluated at admission, as bacterial coinfection more frequently occurs later in the disease evolution.

This study has some strengths. First, all data were collected prospectively with very few missing data for all collected variables. Second, there was an appropriate number of patients in the group that required admission to an ICU, which allowed the study of multiple predictors. Third, the study is reported based on TRIPOD guidelines ^37^

There are limitations to this study. The study included a single-center and very sick population with high rates of ICU admission; therefore, the score may perform well in severe hospitalized patients with COVID-pneumonia. There is still a need for external validation. Another limitation is that medical practice and admission criteria to ICU may vary between institutions and countries, especially in stressed health systems. This score was derived from a population living in Mexico City at >7300 feet over the sea level. The mean partial pressure of oxygen at this altitude has been estimated at around 66mmHg (estimated normal baseline P/F ratio ≈ 314) ^38,39^. Therefore, we used the S/F ratio to account for respiratory compensation and the right-shift of the hemoglobin dissociation curve that takes place at higher altitudes. Finally, the thresholds set to define the low-, moderate- and high-risk groups should be adapted to local needs and ICU availability. Therefore, we provided the estimated percentages of ICU admission for each sum of the score.

In summary, the ABC-GOALS score represents a tool to evaluate patients with COVID-19 at admission to the ED, designed to timely predict their risk of admission to an ICU. This score may help early referral and planning of attention during the COVID-19 pandemic.

## Data Availability

The data included into this manuscript has not yet been subject of peer review.

## FUNDING

None.

## DECLARATION OF INTEREST

None.

## ACKNOWLEDGEMENTS

The authors want to acknowledge all the health care team at Instituto Nacional de Ciencias Médicas y Nutrición Salvador Zubirán at Mexico City for their extraordinary work during this pandemic. We thank Pablo Galindo-Vallejo and Mauricio Moreno-Yañez for the development of the mobile phone application.

---

**Supplementary Table 1.** Characteristics at admission to the emergency department of all the subjects included in the development cohort.

|  | All patients<br>n=329 | Critical Care<br>n=115 | General wards<br>n=214 | p-value |
| --- | --- | --- | --- | --- |
| Age, years | 49 (41-60) | 53 (43-62) | 49 (39-59) | 0.009 |
| Male gender, n (%) | 211 (64) | 88 (77) | 123 (58) | <0.001 |
| Current/past smoker, n (%) | 23 (7) | 10 (9) | 13 (6) | 0.497 |
| **Comorbidities** |  |  |  |  |
| Diabetes mellitus, n (%) | 80 (24) | 37 (32) | 43 (20) | 0.016 |
| Hypertension, n (%) | 88 (27) | 33 (29) | 55 (26) | 0.602 |
| Obesity, n (%) | 132 (40) | 59 (51) | 73 (34) | 0.003 |
| CKD, n (%) | 19 (6) | 7 (6) | 12 (6) | 0.807 |
| None, n (%) | 100 (30) | 18 (16) | 82 (38) | <0.001 |
| Charlson index, points | 1 (0-2) | 1 (0-3) | 1 (0-2) | 0.001 |
| 0 points | 141 (43) | 36 (31) | 105 (49) | 0.002 |
| 1-2 points | 125 (38) | 47 (41) | 78 (36) |  |
| ≥3 points | 63 (19) | 32 (28) | 31 (15) |  |
| **Clinical Presentation** |  |  |  |  |
| Fever, n (%) | 302 (92) | 104 (90) | 198 (93) | 0.532 |
| Cough, n (%) | 285 (87) | 103 (90) | 182 (85) | 0.309 |
| Myalgia / arthralgia, n (%) | 148 (45) | 40 (35) | 108 (51) | 0.007 |
| Sore throat, n (%) | 100 (30) | 36 (31) | 64 (30) | 0.803 |
| Rhinorrhea, n (%) | 51 (16) | 19 (17) | 32 (15) | 0.750 |
| Headache, n (%) | 162 (49) | 56 (49) | 106 (50) | 0.908 |
| Dyspnea, n (%) | 229 (70) | 98 (85) | 131 (61) | <0.001 |
| Diarrhea, n (%) | 56 (17) | 21 (18) | 35 (16) | 0.759 |
| Anosmia, n (%) | 17 (5) | 4 (4) | 13 (6) | 0.435 |
| Days with symptoms | 7 (5-10) | 7 (5-9) | 8 (5-10) | 0.595 |
| **Physical examination** |  |  |  |  |
| Body mass index, kg/m<sup>2</sup> | 29.0 (26.5-31.2) | 29.6 (26.9-33.4) | 28.9 (26.3-30.6) | 0.006 |
| Mean AP, mmHg | 90 (83-98) | 89 (82-97) | 91 (84-99) | 0.152 |
| Systolic AP<100 mmHg | 16 (5) | 11 (10) | 5 (2) | 0.001 |
| Respiratory rate, per min | 24 (21-30) | 28 (24-36) | 24 (20-28) | <0.001 |
| <24 per minute | 122 (37) | 21 (18) | 101 (47) | <0.001 |
| 24-28 per minute | 116 (35) | 37 (32) | 79 (37) |  |
| >28 per minute | 91 (28) | 57 (50) | 34 (16) |  |
| **Laboratory evaluation** |  |  |  |  |
| Total leukocytes, x10<sup>3</sup>/μL | 7.1 (5.3-9.5) | 9.0 (5.8-12.1) | 6.7 (5.1-8.7) | <0.001 |
| Neutrophils, x10<sup>3</sup>/μL | 5.9 (2.8-8.2) | 7.7 (4.8-10.7) | 5.2 (3.6-7.1) | <0.001 |
| Lymphocytes, x10<sup>3</sup>/μL | 0.8 (0.6-1.1) | 0.7 (0.5-1.0) | 0.9 (0.7-1.1) | <0.001 |
| Monocytes, x10<sup>3</sup>/μL | 0.4 (0.3-0.6) | 0.4 (0.3-0.5) | 0.4 (0.3-0.6) | 0.526 |
| Hemoglobin, g/dL | 15.5 (14.4-16.6) | 15.5 (14.4-16.5) | 15.6 (14.2-16.6) | 0.843 |
| Platelets, x10<sup>3</sup> | 208 (165-257) | 215 (166-266) | 206 (165-256) | 0.515 |
| Total bilirubin, mg/dL | 0.55 (0.43-0.75) | 0.62 (0.46-0.84) | 0.53 (0.43-0.69) | 0.005 |
| Direct bilirubin, mg/dL | 0.16 (0.11-0.22) | 0.18 (0.14-0.27) | 0.14 (0.11-0.21) | <0.001 |
| ALT, U/L | 34.3 (23.3-52.7) | 37.3 (23.9-52.7) | 33.7 (22.3-52.6) | 0.574 |
| AST, U/L | 39.3 (27.9-57.6) | 46.6 (33.3-67.8) | 36.5 (26.5-51.2) | <0.001 |
| Alkaline phosphatase, mg/dL | 83 (68-105) | 84 (70-107) | 83 (67-103) | 0.608 |
| Serum albumin, g/dL | 3.8 (3.4-4.1) | 3.5 (3.2-3.9) | 3.9 (2.6-4.1) | <0.001 |
| <3.5 g/dL, n (%) | 98 (30) | 57 (50) | 41 (19) | <0.001 |
| Serum globulins, g/dL | 3.2 (3.0-3.5) | 3.3 (3.0-3.6) | 3.2 (2.9-3.5) | 0.608 |
| Glucose, mg/dL | 115 (100-147) | 138 (111-213) | 107 (97-128) | <0.001 |
| >200 mg/dL, n (%) | 50 (15) | 29 (25) | 21 (10) | <0.001 |
| BUN, mg/dL | 14.0 (10.5-19.8) | 16.6 (12.8-24.0) | 12.7 (9.7-17.9) | <0.001 |
| Creatinine, mg/dL | 0.93 (0.75-1.15) | 1.00 (0.85-1.24) | 0.89 (0.72-1.10) | <0.001 |
| Sodium, mEq/L | 135 (132-137) | 134 (132-138) | 135 (133-137) | 0.200 |
| Potassium, mEq/L | 4.0 (3.7-4.3) | 4.0 (3.6-4.3) | 4.0 (3.7-4.3) | 0.823 |
| Chloride, mEq/L | 101 (98-104) | 100 (97-104) | 102 (99-104) | 0.042 |
| Total serum CO<sub>2</sub>, mmol/L | 23.2 (21.5-25.0) | 22.6 (20.8-24.4) | 23.4 (21.9-25.2) | 0.001 |
| Corrected calcium, mg/dL | 8.6 (8.5-9.0) | 8.6 (8.4-8.9) | 8.7 (8.5-9.0) | <0.001 |
| Phosphorus, mg/dL | 3.2 (2.8-3.7) | 3.3 (2.7-3.9) | 3.2 (2.8-3.6) | 0.114 |
| Magnesium, mg/dL | 2.1 (2.0-2.3) | 2.2 (2.0-2.4) | 2.1 (2.0-2.3) | 0.076 |
| Creatine kinase, U/L | 103 (61-222) | 142 (72-276) | 88 (57-166) | 0.003 |
| Lactate dehydrogenase, U/L | 336 (259-456) | 474 (344-578) | 288 (230-380) | <0.001 |
| > ULN (271 U/L), n (%) | 230 (70) | 106 (92) | 124 (58) | <0.001 |
| C reactive protein, mg/dL | 12.6 (5.4-19.3) | 18.3 (12.6-25.7) | 7.8 (3.4-15.6) | <0.001 |
| Ferritin, ng/mL | 506 (254-939) | 740 (423-1092) | 375 (207-784) | <0.001 |
| Troponin I, pg/mL | 4.6 (2.9-8.5) | 7.8 (4.9-19.4) | 3.7 (2.6-5.6) | <0.001 |
| >ULN (20pg/mL), n (%) | 35 (10.6) | 28 (24.3) | 7 (3.3) | <0.001 |
| Fibrinogen, mg/dL | 604 (457-723) | 688 (542-807) | 499 (434-678) | <0.001 |
| D-dimer, ng/mL | 628 (398-1072) | 761 (495-1323) | 523 (358-958) | <0.001 |
| **Arterial blood gas analysis** |  |  |  |  |
| pH | 7.44 (7.42-7.47) | 7.44 (7.39-7.46) | 7.45 (7.42-7.47) | <0.001 |
| Bicarbonate, mmol/L | 21.5 (19.9-22.9) | 21.0 (18.6-23.5) | 21.6 (20.5-22.8) | 0.122 |
| Lactate, mmol/L | 1.2 (0.9-1.6) | 1.5 (1.0-2.1) | 1.1 (0.9-1.4) | <0.001 |
| PaO<sub>2</sub>/FiO<sub>2</sub> ratio | 247 (175-291) | 177 (107-247) | 269 (229-305) | <0.001 |
| SaO<sub>2</sub>/FiO<sub>2</sub> ratio | 330 (211-426) | 193 (149-325) | 410 (315-433) | <0.001 |
| <300, n (%) | 124 (38) | 80 (70) | 44 (21) | <0.001 |
| **Scores at presentation** |  |  |  |  |
| NEWS-2, points | 7 (5-8) | 8 (7-10) | 6 (4-7) | <0.001 |
| SOFA - PaO<sub>2</sub>/FiO<sub>2</sub> | 2 (2-3) | 3 (2-4) | 2 (2-3) | <0.001 |
| SOFA - SaO<sub>2</sub>/FiO<sub>2</sub> | 1 (0-2) | 2 (1-3) | 0 (0-1) | <0.001 |
| COVID-GRAM score, points | 116 (98-137) | 135 (115-156) | 103 (87-125) | <0.001 |
| CALL score, points | 9 (7-11) | 10 (8-11) | 8 (7-10) | <0.001 |
| CURB-65 score, points | 0 (0-1) | 1 (0-2) | 0 (0-1) | <0.001 |
| MuLBSTA score, points | 9 (5-11) | 9 (5-11) | 7 (5-9) | <0.001 |
| ROX index | 13.6 (7.1-18.6) | 6.9 (4.5-12.9) | 16.4 (12.3-20.7) | <0.001 |
| **Chest CT scan** |  |  |  |  |
| Multi-lobar, n (%) | 329 (100) | 115 (100) | 214 (100) | 1.000 |
| Extension, n (%) |  |  |  |  |
| Mild (≤20%) | 57 (17) | 3 (3) | 54 (25) | <0.001 |
| Moderate (21-50%) | 141 (43) | 26 (23) | 115 (54) |  |
| Severe (>50%) | 131 (40) | 86 (75) | 45 (21) |  |

**Abbreviations.** CKD, chronic kidney disease; AP, arterial pressure; ALT, alanine aminotransferase; AST, aspartate aminotransferase; BUN, blood urea nitrogen; CO<sub>2</sub>, total diluted carbon dioxide; PaO<sub>2</sub>/FiO<sub>2</sub> ratio, ratio of arterial pressure of oxygen to fraction of inspired oxygen; SaFiO<sub>2</sub>/FiO<sub>2</sub>, ratio of the percentage of hemoglobin saturation by oxygen to fraction of inspired oxygen; NEWS-2, National Early Warning Score 2; SOFA – PaO<sub>2</sub>/FiO<sub>2</sub>, Sequential Organ Failure Assessment score with respiratory points obtained by PaO<sub>2</sub>/FiO<sub>2</sub> ratio; SOFA-SaO<sub>2</sub>/FiO<sub>2</sub>, Sequential Organ Failure Assessment score with respiratory points obtained by SaO<sub>2</sub>/FiO<sub>2</sub>; MuLBSTA, Multi-lobar infiltrate, Lymphocyte count, Bacterial co-infection, Smoking history, hypertension, Age>65 years score for mortality from viral pneumonia; ROX index, S/F ratio divided by the respiratory rate to predict the risk of intubation in hypoxemic respiratory failure ULN, upper limit of normal.

**Supplementary Table 2.** Variables explored by bivariate logistic regression analysis.

|  | OR | 95% CI | p-value |
| --- | --- | --- | --- |
| **Demographics variables** |  |  |  |
| Age, per year | 1.021 | 1.004-1.038 | 0.015* |
| Age ≥50 years, vs. less | 1.716 | 1.085-2.713 | 0.021* |
| Male gender, vs. female | 2.411 | 1.449-4.013 | 0.001* |
| **Previous medical history** |  |  |  |
| Current or previous smoker, vs. no | 1.473 | 0.625-3.471 | 0.376 |
| Hypertension, vs. no | 1.163 | 0.701-1.932 | 0.559 |
| Diabetes mellitus, vs. no | 1.886 | 1.128-3.156 | 0.016* |
| Chronic kidney disease, vs. no | 1.122 | 0.429-2.935 | 0.814 |
| Obesity (BMI>30kg/m<sup>2</sup>), vs. no | 2.035 | 1.282-3.230 | 0.003* |
| No comorbidities, vs. yes | 0.299 | 0.168-0.530 | 0.000* |
| Charlson index, per point | 1.235 | 1.071-1.425 | 0.004* |
| Charlson index categories |  |  |  |
| 0 points | 1.000 | reference | reference |
| 1-2 points | 1.757 | 1.041-2.967 | 0.035 |
| ≥ 3 points | 3.011 | 1.616-5.610 | 0.001 |
| **Clinical presentation variables** |  |  |  |
| Days with symptoms, per day | 1.010 | 0.956- 1.066 | 0.729 |
| Three or more days with symptoms | 3.859 | 0.857-17.38 | 0.079 |
| Fever, vs. none | 0.764 | 0.342-1.706 | 0.511 |
| Cough, vs. none | 1.509 | 0.745-3.058 | 0.253* |
| Fatigue/malaise, vs. none | 0.907 | 0.576-1.428 | 0.673 |
| Mialgia/arthralgia, vs. none | 0.523 | 0.328-0.836 | 0.007* |
| Sore throat, vs. none | 1.068 | 0.654-1.745 | 0.793 |
| Rinorrhea, vs. none | 1.126 | 0.606-2.091 | 0.708 |
| Headache, vs. none | 0.967 | 0.615-1.522 | 0.885 |
| Dyspnea, vs. none | 3.652 | 2.037-6.548 | 0.000* |
| Diarrhea, vs. none | 1.143 | 0.630-2.073 | 0.661 |
| Anosmia, vs. none | 0.557 | 0.177-1.750 | 0.316 |
| **Physical examination variables** |  |  |  |
| Mean arterial pressure, per mmHg | 0.989 | 0.971-1.007 | 0.233 |
| MAP<65mmHg, vs. higher | 3.770 | 0.338-42.03 | 0.281 |
| Systolic arterial pressure, per mmHg | 0.991 | 0.978-1.004 | 0.184 |
| SAP<100mmHg, vs. higher | 4.421 | 1.497-13.06 | 0.007* |
| Diastolic arterial pressure, per mmHg | 0.991 | 0.973-1.010 | 0.369 |
| Respiratory rate, per 1x' | 1.137 | 1.094-1.183 | 0.000* |
| Respiratory rate |  |  |  |
| <24 per minute | 1.000 | reference | reference |
| 24-28 per minute | 2.253 | 1.223-4.150 | 0.009 |
| >28 per minute | 8.063 | 4.279-15.19 | 0.000* |
| Body mass index, per kg/m<sup>2</sup> | 1.080 | 1.030-1.132 | 0.001* |
| **Laboratory variables** |  |  |  |
| Total leukocytes, per 1x10<sup>3</sup> | 1.209 | 1.127-1.296 | 0.000* |
| Neutrophils, per 1x10<sup>3</sup> | 1.000 | 1.000-1.000 | 0.000* |
| Lymphocytes, per 1x10<sup>3</sup> | 0.999 | 0.998-1.000 | 0.001* |
| Monocytes, per 1x10<sup>3</sup> | 1.001 | 0.999-1.001 | 0.596 |
| Hemoglobin, per g/dL | 0.973 | 0.860-1.101 | 0.666 |
| Platelets, per 1x10<sup>6</sup> | 1.001 | 0.998-1.004 | 0.559 |
| Total bilirubin, per mg/dL | 1.753 | 0.999-3.076 | 0.050 |
| Direct bilirubin, per mg/dL | 0.963 | 0.806-1.151 | 0.681 |
| ALT, per U/L | 1.002 | 0.999-1.005 | 0.256 |
| AST, per U/L | 1.005 | 1.000-1.010 | 0.033* |
| AST ≥40U/L, vs. lower | 2.310 | 1.452-3.676 | 0.000* |
| Alkaline phosphatase, per mg/dL | 1.001 | 0.996-1.006 | 0.773 |
| Serum albumin, per mg/dL | 0.166 | 0.094-0.294 | 0.000* |
| Serum albumin <3.5mg/dL, vs. higher | 4.147 | 2.516-6.834 | 0.000* |
| Serum globulins, per mg/dL | 1.341 | 0.812-2.215 | 0.252 |
| Glucose, per mg/dL | 1.008 | 1.005-1.012 | 0.000* |
| Glucose > 200 mg/dL, vs. lower | 3.099 | 1.673-5.740 | 0.000* |
| Blood urea nitrogen, per mg/dL | 1.048 | 1.023-1.073 | 0.000* |
| Serum creatinine, per mg/dL | 1.638 | 1.003-2.676 | 0.048* |
| Sodium, per mEq/L | 0.967 | 0.914-1.024 | 0.253 |
| Potassium, per mEq/L | 1.124 | 0.853-1.481 | 0.405 |
| Chloride, per mEq/L | 0.945 | 0.894-0.997 | 0.040* |
| Total CO<sub>2</sub>, per mmol/L | 0.873 | 0.803-0.948 | 0.001* |
| Corrected calcium, per mg/dL | 1.008 | 0.851-1.194 | 0.927 |
| Phosphorus, per mg/dL | 1.452 | 1.091-1.931 | 0.011* |
| Magnesium per mg/dL | 1.821 | 0.875-3.793 | 0.109 |
| Creatine kinase, per U/L | 1.001 | 1.000-1.002 | 0.067 |
| Lactate dehydrogenase, per U/L | 1.006 | 1.004-1.07 | 0.000* |
| ≥ upper limit of normal, vs. lower | 8.548 | 4.109-17.79 | 0.000* |
| C-reactive protein, per mg/dL | 1.114 | 1.082-1.148 | 0.000* |
| ≥5mg/dL, vs. lower | 29.252 | 7.026-121.8 | 0.000* |
| ≥10mg/dL, vs. lower | 7.942 | 4.438-14.215 | 0.000* |
| Ferritin, per ng/mL | 1.001 | 1.000-1.001 | 0.000* |
| ≥400ng/mL, vs. lower | 3.579 | 2.154-5.947 | 0.000* |
| Troponin I, per pg/mL | 1.006 | 1.000-1.012 | 0.047* |
| ≥upper limit of normal, vs. lower | 9.471 | 3.987-22.501 | 0.000* |
| Fibrinogen, per mg/dL | 1.004 | 1.002-1.005 | 0.000* |
| ≥500ng/mL, vs. lower | 3.249 | 1.963-5.376 | 0.000* |
| D dimer, per ng/mL | 1.000 | 1.000-1.000 | 0.036* |
| ≥500ng/mL, vs. lower | 2.620 | 1.589-4.318 | 0.000* |
| **Arterial blood gas analysis** |  |  |  |
| pH, per unit | 0.000 | 0.000-0.003 | 0.000* |
| Bicarbonate, per mmol/L | 0.948 | 0.880-1.021 | 0.948 |
| Lactate, per mmol/L | 2.352 | 1.620-3.415 | 0.000* |
| P/F ratio, per unit | 0.989 | 0.985-0.992 | 0.000* |
| S/F ratio, per unit | 0.990 | 0.988-0.992 | 0.000* |
| S/F ratio<300, vs. higher | 8.831 | 5.264-14.815 | 0.000* |
| **Scores** |  |  |  |
| NEWS-2, per point | 1.604 | 1.409-1.826 | 0.000* |
| SOFA – PaO<sub>2</sub>/FiO<sub>2</sub>, per point | 1.911 | 1.551-2.355 | 0.000* |
| SOFA – SaO<sub>2</sub>/FiO<sub>2</sub>, per point | 2.090 | 1.711-2.551 | 0.000* |
| COVID-GRAM score, per point | 1.037 | 1.027-1.047 | 0.000* |
| CALL score, per point | 1.483 | 1.311-1.677 | 0.000* |
| CURB-65, per point | 2.409 | 1.831-3.170 | 0.000* |
| ROX index, per point | 0.822 | 0.785-0.862 | 0.000* |
| MuLBSTA score, per point | 1.200 | 1.099-1.310 | 0.000* |

| Chest CT scan |  |  |  |
| --- | --- | --- | --- |
| Pneumonia extension |  |  |  |
| Mild-moderate (<50%) | 1.000 | reference | Reference |
| Severe (>50%) | 11.137 | 6.529-18.99 | <0.001 |

**Abbreviations.** BMI, body mass index; ALT, alanine aminotransferase; AST, aspartate aminotransferase; P/F ratio, ratio of arterial pressure of oxygen to fraction of inspired oxygen; S/F ratio, ratio of oxygen hemoglobin saturation to fraction of inspired oxygen; NEWS-2, National Early Warning Score 2; SOFA – PaO<sub>2</sub>/FiO<sub>2</sub>, Sequential Organ Failure Assessment score with respiratory points obtained by PaO<sub>2</sub>/FiO<sub>2</sub> ratio; SOFA-SaO<sub>2</sub>/FiO<sub>2</sub>, Sequential Organ Failure Assessment score with respiratory points obtained by SaO<sub>2</sub>/FiO<sub>2</sub>, MuLBSTA, Multi-lobar infiltrate, Lymphocyte count, Bacterial co-infection, Smoking history, hypertension, Age>65 years score for mortality from viral pneumonia; ROX index, S/F ratio divided by the respiratory rate to predict the risk of intubation in hypoxemic respiratory failure.

**Supplementary Table 3.** Characteristics at admission to the emergency department of all the subjects included in the validation cohort.

|  | All patients<br>n=240 | Critical Care<br>n= 115 | General wards<br>n= 125 | p-value |
| --- | --- | --- | --- | --- |
| Age, years | 52 (43-60) | 55 (46-65) | 50 (42-58) | 0.001 |
| Male gender, n (%) | 166 (69) | 86 (75) | 80 (74) | 0.071 |
| Current/past smoker, n (%) | 36 (15) | 13 (11) | 23 (18) | 0.124 |
| **Comorbidities** |  |  |  |  |
| Diabetes mellitus, n (%) | 78 (33) | 47 (41) | 31 (25) | 0.008 |
| Hypertension, n (%) | 74 (31) | 39 (34) | 35 (28) | 0.322 |
| Obesity, n (%) | 127 (53) | 64 (56) | 63 (50) | 0.415 |
| CKD, n (%) | 12 (5) | 5 (4) | 7 (6) | 0.657 |
| None, n (%) | 50 (21) | 19 (17) | 31 (25) | 0.115 |
| Charlson index, points |  |  |  |  |
| 0 points | 80 (33) | 29 (25) | 51 (41) | <0.001 |
| 1-2 points | 11 (46) | 48 (42) | 63 (50) |  |
| ≥3 points | 49 (21) | 38 (33) | 11 (9) |  |
| **Clinical Presentation** |  |  |  |  |
| Fever, n (%) | 201 (84) | 94 (82) | 107 (86) | 0.418 |
| Cough, n (%) | 191 (80) | 94 (82) | 97 (78) | 0.427 |
| Myalgia / arthralgia, n (%) | 145 (60) | 23 (20) | 48 (38) | 0.002 |
| Sore throat, n (%) | 55 (23) | 27 (24) | 28 (22) | 0.843 |
| Rhinorrhea, n (%) | 32 (13) | 16 (14) | 16 (13) | 0.800 |
| Headache, n (%) | 121 (50) | 52 (45) | 69 (55) | 0.122 |
| Dyspnea, n (%) | 211 (88) | 108 (94) | 103 (82) | 0.006 |
| Diarrhea, n (%) | 42 (18) | 13 (11) | 29 (23) | 0.015 |
| Anosmia, n (%) | 18 (8) | 7 (6) | 11 (9) | 0.425 |
| Days with symptoms | 8 (5-10) | 7 (5-10) | 8 (5-11) | 0.431 |
| **Physical examination** |  |  |  |  |
| Body mass index, kg/m<sup>2</sup> | 30.1 (26.7-33.9) | 30.1 (26.6-34.4) | 29.8 (26.8-33.8) | <0.001 |
| Mean AP, mmHg | 92 (83-97) | 90 (78-97) | 93 (87-100) | 0.011 |
| Systolic AP<100 mmHg | 10 (4) | 9 (8) | 1 (1) | <0.001 |
| Respiratory rate, per min |  |  |  |  |
| <24 per minute | 48 (20) | 10 (9) | 38 (30) | <0.001 |
| 24-28 per minute | 65 (27) | 24 (21) | 41 (33) |  |
| >28 per minute | 127 (53) | 81 (70) | 46 (37) |  |
| **Laboratory evaluation** |  |  |  |  |
| Total leukocytes, x10<sup>3</sup>/μL | 8.6 (6.5-12.7) | 9.9 (7.2-14.4) | 8.0 (5.7-10.7) | <0.001 |
| Neutrophils, x10<sup>3</sup>/μL | 7.3 (5.4-10.9) | 8.2 (6.2-13.1) | 6.4 (4.8-8.8) | <0.001 |
| Lymphocytes, x10<sup>3</sup>/μL | 0.8 (0.5-1.0) | 0.7 (0.5-1.0) | 0.8 (0.6-1.1) | 0.018 |
| Monocytes, x10<sup>3</sup>/μL | 0.4 (0.3-0.6) | 0.4 (0.3-0.6) | 0.4 (0.3-0.6) | 0.795 |
| Hemoglobin, g/dL | 15.5 (14.3-16.3) | 15.1 (14.2-16.3) | 15.7 (14.4-16.4) | 0.255 |
| Platelets, x10<sup>3</sup> | 235 (182-307) | 239 (190-307) | 234 (175-308) | 0.447 |
| Total bilirubin, mg/dL | 0.6 (0.5-0.9) | 0.7 (0.5-1.0) | 0.6 (0.5-0.8) | 0.038 |
| Direct bilirubin, mg/dL | 0.2 (0.1-0.3) | 0.2 (0.1-0.3) | 0.2 (0.1-0.2) | 0.008 |
| ALT, U/L | 38 (25-60) | 44 (27-62) | 34 (23-52) | 0.008 |
| AST, U/L | 44 (32-64) | 49 (39-65) | 40 (27-60) | 0.001 |
| Alkaline phosphatase, mg/dL | 93 (73-120) | 97 (76-131) | 87 (71-112) | 0.068 |
| Serum albumin, g/dL | 3.6 (3.3-3.9) | 3.4 (3.1-3.7) | 3.8 (3.5-4.0) | <0.001 |
| <3.5 g/dL, n (%) | 103 (43) | 69 (60) | 34 (27) | <0.001 |
| Serum globulins, g/dL | 3.4 (3.1-3.6) | 3.4 (3.1-3.6) | 3.4 (3.2-3.7) | 0.991 |
| Glucose, mg/dL | 132 (107-182) | 153 (118-230) | 119 (104-145) | <0.001 |
| >200 mg/dL, n (%) | 51 (21) | 36 (31) | 15 (12) | <0.001 |
| BUN, mg/dL | 18 (12-26) | 21 (15-30) | 15 (11-19) | <0.001 |
| Creatinine, mg/dL | 1.0 (0.8-1.2) | 1.0 (0.8-1.3) | 0.9 (0.8-1.1) | 0.006 |
| Sodium, mEq/L | 135 (133-138) | 134 (132-137) | 135 (133-138) | 0.038 |
| Potassium, mEq/L | 4.1 (3.8-4.5) | 4.1 (3.9-4.6) | 4.1 (3.8-4.4) | 0.166 |
| Chloride, mEq/L | 101 (98-103) | 101 (97-104) | 101 (98-103) | 0.475 |
| Total serum CO<sub>2</sub>, mmol/L | 22.8 (20.8-24.7) | 22.1 (19.8-24.4) | 23.3 (21.2-25.6) | 0.001 |
| Corrected calcium, mg/dL | 8.7 (8.5-9.0) | 8.7 (8.5-9.0) | 8.7 (8.5-9.0) | 0.879 |
| Phosphorus, mg/dL | 3.3 (2.9-3.8) | 3.3 (2.9-3.9) | 3.2 (2.9-3.7) | 0.453 |
| Magnesium, mg/dL | 2.2 (2.0-2.4) | 2.3 (2.1-2.4) | 2.2 (2.0-2.4) | 0.028 |
| Creatine kinase, U/L | 109 (60-242) | 143 (72-242) | 96 (59-243) | 0.011 |
| Lactate dehydrogenase, U/L | 413 (317-548) | 518 (403-586) | 342 (277-429) | <0.001 |
| > ULN (271 U/L), n (%) | 206 (86) | 110 (96) | 96 (77) | <0.001 |
| C reactive protein, mg/dL | 17.3 (10.7-25.7) | 20.0 (13.8-29.3) | 14.3 (9.0-20.0) | <0.001 |
| Ferritin, ng/mL | 747 (399-1134) | 812 (514-1275) | 712 (345-1056) | 0.006 |
| Troponin I, pg/mL | 6.2 (3.8-14.4) | 10.5 (5.4-49.2) | 4.6 (3.0-7.2) | <0.001 |
| >ULN (20pg/mL), n (%) | 51 (21) | 39 (34) | 12 (10) | <0.001 |
| Fibrinogen, mg/dL | 735 (616-887) | 770 (648-934) | 703 (554-858) | 0.002 |
| D-dimer, ng/mL | 849 (525-1396) | 1088 (661-1967) | 715 (437-1092) | <0.001 |
| **Arterial blood gas analysis** |  |  |  |  |
| pH | 7.43 (7.40-7.47) | 7.42 (7.38-7.46) | 7.44 (7.42-7.47) | 0.001 |
| Bicarbonate, mmol/L | 21.4 (19.6-23.1) | 21 (19-23) | 22 (20-23) | 0.014 |
| Lactate, mmol/L | 1.5 (1.2-2.1) | 1.8 (1.4-2.5) | 1.3 (1.0-1.7) | <0.001 |
| PaO<sub>2</sub>/FiO<sub>2</sub> ratio | 197 (122-260) | 133 (98-199) | 241 (193-279) | <0.001 |
| SaO<sub>2</sub>/FiO<sub>2</sub> ratio | 258 (158-371) | 159 (148-252) | 330 (252-422) | <0.001 |
| <300, n (%) | 142 (59) | 99 (86) | 43 (34) | <0.001 |
| **Scores at presentation** |  |  |  |  |
| NEWS-2, points | 8 (6-9) | 8 (7-9) | 7 (6-9) | <0.001 |
| SOFA - PaO<sub>2</sub>/FiO<sub>2</sub> | 3 (2-4) | 4 (3-4) | 3 (2-3) | <0.001 |
| SOFA - SaO<sub>2</sub>/FiO<sub>2</sub> | 2 (0-3) | 2 (2-3) | 1 (0-2) | <0.001 |
| COVID-GRAM score, points | 129 (113-149) | 140 (126-162) | 118 (104-134) | <0.001 |
| CALL score, points | 10 (8-11) | 10 (9-11) | 9 (8-10) | <0.001 |
| CURB-65 score, points | 1 (0-2) | 2 (1-2) | 1 (0-1) | <0.001 |
| MuLBSTA score, points | 9 (7-11) | 9 (7-11) | 9 (5-11) | <0.001 |
| ROX index | 8.6 (4.9-13.6) | 5.4 (4.0-8.1) | 13.1 (8.5-17.2) | <0.001 |
| **Chest CT scan** |  |  |  |  |
| Multi-lobar, n (%) | 240 (100) | 115 (100) | 125 (100) | 1.000 |
| Extension, n (%) |  |  |  |  |
| Mild (≤20%) | 6 (3) | 0 (0) | 6 (5) | <0.001 |
| Moderate (21-50%) | 89 (37) | 23 (20) | 66 (53) |  |
| Severe (>50%) | 145 (60) | 92 (80) | 53 (42) |  |

**Abbreviations.** CKD, chronic kidney disease; AP, arterial pressure; ALT, alanine aminotransferase; AST, aspartate aminotransferase; BUN, blood urea nitrogen; CO<sub>2</sub>, total diluted carbon dioxide; PaO<sub>2</sub>/FiO<sub>2</sub> ratio, ratio of arterial pressure of oxygen to fraction of inspired oxygen; SaFiO<sub>2</sub>/FiO<sub>2</sub>, ratio of the percentage of hemoglobin saturation by oxygen to fraction of inspired oxygen; NEWS-2, National Early Warning Score 2; SOFA – PaO<sub>2</sub>/FiO<sub>2</sub>, Sequential Organ Failure Assessment score with respiratory points obtained by PaO<sub>2</sub>/FiO<sub>2</sub> ratio; SOFA-SaO<sub>2</sub>/FiO<sub>2</sub>, Sequential Organ Failure Assessment score with respiratory points obtained by SaO<sub>2</sub>/FiO<sub>2</sub>; MuLBSTA, Multi-lobar infiltrate, Lymphocyte count, Bacterial co-infection, Smoking history, hypertension, Age>65 years score for mortality from viral pneumonia; ROX index, S/F ratio divided by the respiratory rate to predict the risk of intubation in hypoxemic respiratory failure ULN, upper limit of normal.

**Supplementary Figure 1.**
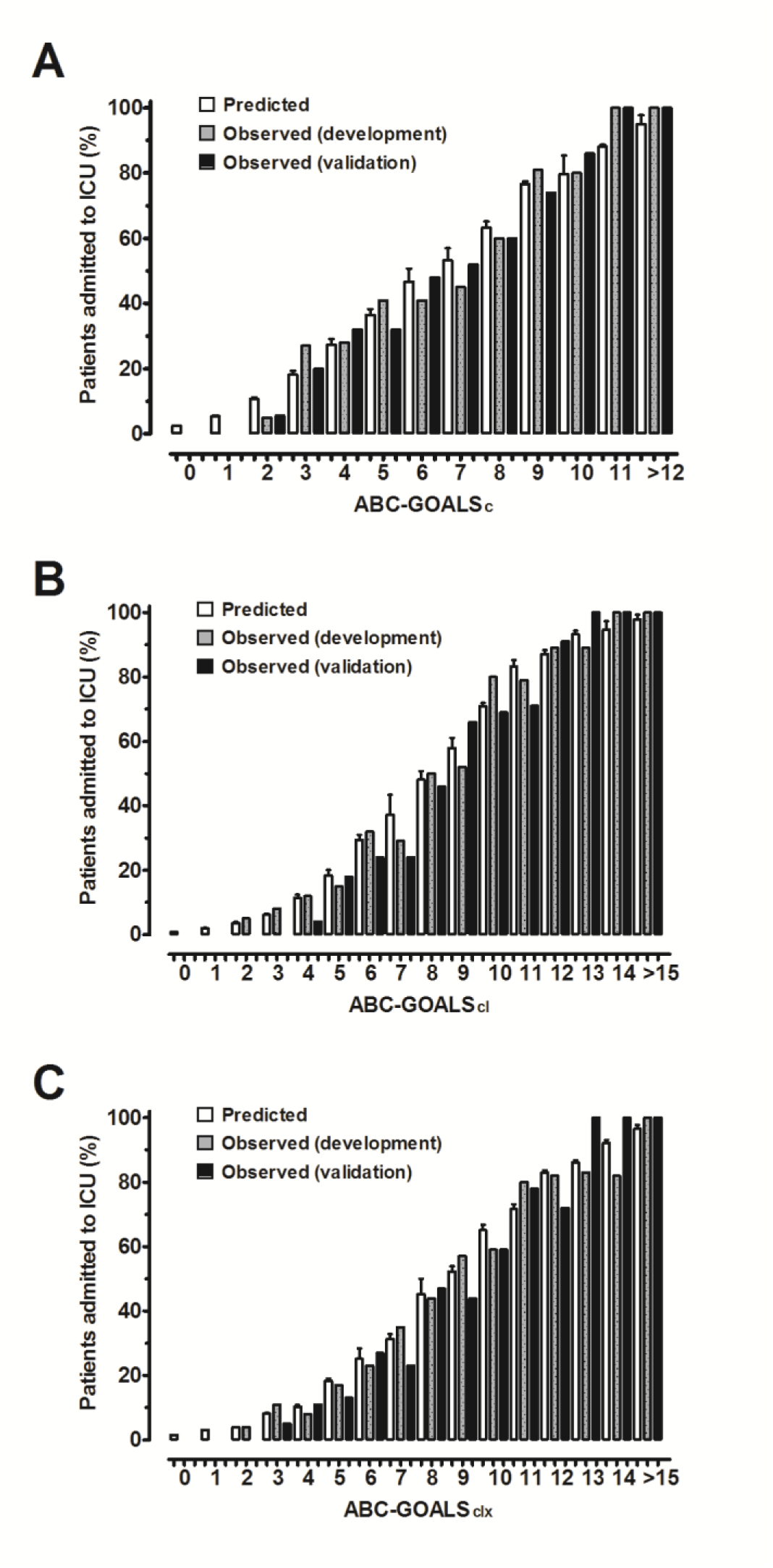
Predicted and observed percentages of patients who required admission to ICU by each point of increment in the ABC-GOALS_c_ (A), ABC-GOALS_cl_ (B) and ABC-GOALS_clx_ (C) scores.

## Notes

### Author Declarations

Instituto Nacional de Ciencias Medicas y Nutricion Salvador Zubiran research and ethics boards.

